# Long-lasting cellular immunity to SARS-CoV-2 following infection or vaccination and implications for booster strategies

**DOI:** 10.1101/2021.12.29.21268469

**Authors:** Alessio Mazzoni, Anna Vanni, Michele Spinicci, Giulia Lamacchia, Seble Tekle Kiros, Arianna Rocca, Manuela Capone, Nicoletta Di Lauria, Lorenzo Salvati, Alberto Carnasciali, Elisabetta Mantengoli, Parham Farahvachi, Lorenzo Zammarchi, Filippo Lagi, Maria Grazia Colao, Francesco Liotta, Lorenzo Cosmi, Laura Maggi, Alessandro Bartoloni, Gian Maria Rossolini, Francesco Annunziato

**Affiliations:** Department of Experimental and Clinical Medicine, University of Florence, Florence, Italy; Infectious and Tropical Diseases Unit, Careggi University Hospital, Florence, Italy; Microbiology and Virology Unit, Careggi University Hospital, Florence, Italy; Immunology and Cell Therapy Unit, Careggi University Hospital, Florence, Italy; Flow cytometry diagnostic center and immunotherapy, Careggi University Hospital, Florence, Italy

## Abstract

Immunization against SARS-CoV-2, the causative agent of coronavirus disease-19 (COVID-19) occurs via natural infection or vaccination. However, it is currently unknown how long infection- or vaccination-induced immunological memory will last. We performed a longitudinal evaluation of immunological memory to SARS-CoV-2 following mRNA vaccination in naïve and COVID-19 recovered individuals. We found that cellular immunity is still detectable 8 months after vaccination, while antibody levels decline significantly especially in naïve subjects. We also found that a booster injection is more efficacious in reactivating immunological memory to spike protein in naïve than in previously SARS-CoV-2 infected subjects. Finally, we observed a similar kinetics of decay of humoral and cellular immunity to SARS-CoV-2 up to one year following natural infection in a cohort of unvaccinated individuals. Short-term persistence of humoral immunity may account for reinfections and breakthrough infections, although long-lived memory B and CD4+ T cells may protect from severe disease. A booster dose restores optimal anti-spike immunity in naïve subjects, while the need for vaccinated COVID-19 recovered subjects has yet to be defined.

## Introduction

Immunization against SARS-CoV-2, the causative agent of coronavirus disease-19 (COVID-19) occurs via natural infection or vaccination. As of December 20^th^, 2021 more than 273 million people have been infected worldwide, with more than 5 million deaths [1]. At least one vaccine dose has been administered to more than 4 billion people, while 3.49 billion people have been fully vaccinated, with considerable differences between high- and low-income countries [1]. In order to contain COVID-19 pandemic it is mandatory to achieve high immunization levels worldwide. However, it is currently unknown how long infection-or vaccination-induced immunological memory will last. Regarding natural infection, it has been shown that although antibody levels decline, memory B and T cells can be detected up to 8 months after recovery in most subjects [2], although 10-20% of them display a weak immunological memory [2,3]. The magnitude of immunological memory to SARS-CoV-2 directly correlates to disease severity, as asymptomatic individuals show reduced levels of virus-specific immunity both in the acute and memory phase [4,5]. Information about longevity of vaccine-induced immunological memory are even scarcer, as the immunization campaign started less than one year ago. Real-world studies have shown that 6 months following the complete mRNA vaccination cycle, protection against COVID-19 is significantly reduced in all age groups, although protection from severe disease and hospitalization remains high [6-9]. This phenomenon may be the result of progressive waning of immune protection. Nonetheless, the occurrence of SARS-CoV-2 variants like Delta (B.1.617.2), and more recently Omicron (B.1.1.529), that show increased transmission capability and immune evasion potential, plays also a relevant role. These data raised the question about the need for a booster dose administration, and many countries are now recommending a third vaccine injection 6 months following the second dose. However, it is still not clear if a third injection is required in all individuals, or if it should be reserved to specific groups based on higher professional exposure risk or increased susceptibility to COVID-19 due to concomitant diseases and corresponding treatment regimens. To further complicate this scenario, it has been shown that the second mRNA vaccine injection is dispensable in subjects with previous SARS-CoV-2 infection, since the first administration is sufficient to maximize their immune response to spike [3,10]. Vaccine delivery to recovered COVID-19 individuals results in the so-called “hybrid immunity”, characterized by increased strength of humoral and cellular response and increased breadth of antibody repertoire than both natural infection and vaccination in naïve individuals [11].

In this work, we performed a longitudinal evaluation of immunological memory to SARS-CoV-2 up to 8 months following mRNA vaccination in naïve and COVID-19 recovered individuals. We also assessed the immunological effects of a booster injection in a cohort of individuals from both groups. Our results show that, despite a progressive decline affecting mainly antibody levels, humoral and cellular responses are still detectable in naïve and recovered COVID-19 individuals 8 months after vaccination. Moreover, we observed that a booster injection is more efficacious in reactivating immunological memory to spike in naïve than in previously SARS-CoV-2 infected subjects. Finally, we also assessed the longevity of immunological memory to SARS-CoV-2 following natural infection up to one year in a cohort of unvaccinated individuals. Also in this case, despite a decline in antibody levels, B and T cells memory responses were still detectable 12 months after infection.

## Results

### Anti-spike antibodies decline more rapidly following vaccination in naïve than in COVID-19 recovered subjects

This is a follow up study of our previous work in which we monitored the vaccine-induced humoral and cellular immune response against SARS-CoV-2 spike protein up to one week after the second vaccine injection in naïve and recovered COVID-19 individuals [3]. Here, in order to evaluate the duration of immunological memory induced by vaccination, we extended our cohort to 15 subjects per group who were longitudinally evaluated up to 8 months after vaccination.

Demographic and clinical characteristics of the 30 recruited subjects are detailed in Supplementary Table S1. We obtained a blood draw at basal time (baseline, before injection of the first dose), after 21 days (before injection of the second dose), then after 1, 6 and 8 months to monitor humoral response and the presence of SARS-CoV-2 spike-specific B and T cells.

As shown in Figure 1A, serum titers of anti-S IgM increased in naïve subjects (left panel) following the first vaccine administration, while exhibited significant inter-individual differences among recovered COVID-19 individuals (middle panel). In both groups, anti-S IgM declined from month 1, reaching values below the cut-off limit at months 6 and 8 (right panel). Anti-S IgA and anti-S IgG peaked in naïve subjects at month 1 after second dose administration, while in recovered COVID-19 subjects they reached the highest levels already after the first injection and remained stable up to month 1 (Fig.1B-C left and middle panels). Of note, these antibodies showed a mild decrease in SARS-CoV-2 experienced subjects at months 6 and 8, while in naïve individuals we observed a significant drop at month 6. However, anti-S IgA and IgG at month 8 displayed significant higher levels than before vaccination in both groups. Anti-RBD IgG increased up to month 1, dropped significantly at month 6 but remained at higher levels than baseline at month 8 in both groups (Fig1D left and middle panels). It should be noted that at all time points of analysis recovered COVID-19 subjects showed significantly higher levels of anti-S IgA and IgG and anti-RBD IgG than naïve individuals (Fig.1B-D right panels). Neutralizing antibodies increased in naïve subjects up to month 1 after vaccination, while they reached the highest level in recovered COVID-19 individuals already after the first injection, remaining stable up to month 1 (Fig.1E left and middle panels). Despite a significant decrease at month 6, neutralizing antibodies declined less sharply than anti-RBD IgG in both groups (Fig.1E right panel). Interestingly, neutralizing antibodies at month 8 were significantly higher than before vaccination in naïve individuals, while we observed no significant difference in the recovered COVID-19 group. Also in this case, recovered COVID-19 individuals showed higher antibody levels than naïve subjects at all the analysed time points.

**Figure 1.**
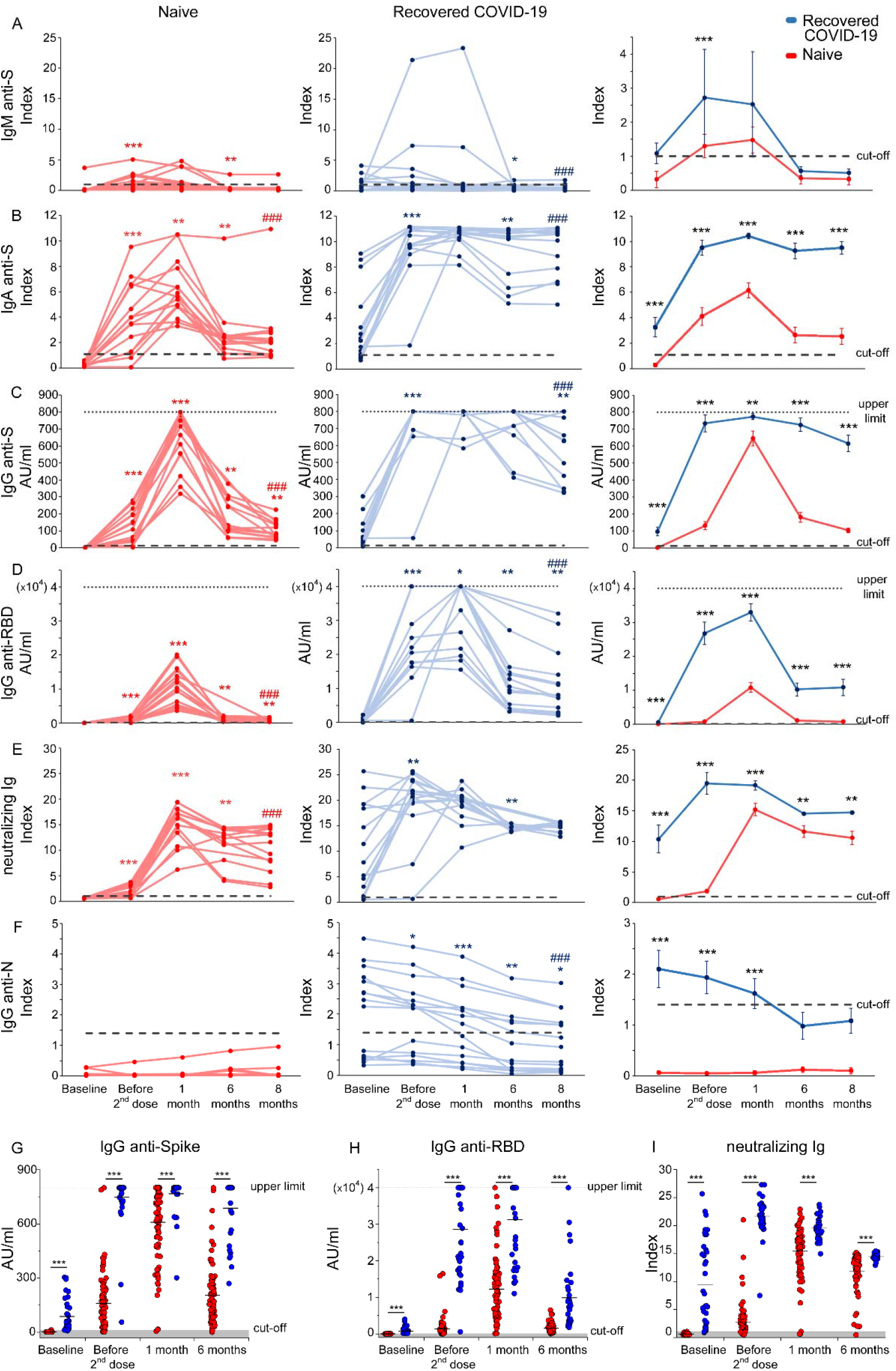
Longitudinal evaluation of anti-SARS-CoV-2 antibody levels in naïve and COVID-19 recovered vaccinated subjects. Evaluation of anti-Spike IgM (A), anti-S1 IgA (B), anti-S IgG (C), anti-RBD IgG (D), S-neutralizing Ig (E), anti-N IgG (F) in naïve and COVID-19 recovered subjects before the first injection, before the second injection and after 1, 6 and 8 months after the complete vaccination cycle. Left column (red lines) represents individual data from 15 naïve subjects, middle column (blue lines) represents individual data from 15 recovered COVID-19 individuals, right column represents mean values ±SE. Dashed lines represent cut-off values, dotted lines are upper detection limits. Red and blue asterisks (left and middle column) refer to paired statistics within each study group compared to the previous time point in the kinetics calculated with Wilcoxon-Signed Rank test. Red and blue hashtag in left and middle columns refer to paired statistics within each study group between baseline and 8 months, calculated with Wilcoxon-Signed Rank test. Black asterisks in the right column represent unpaired statistic between naïve and recovered COVID-19 subjects at each time point calculated with Mann-Whitney test. Evaluation of anti-Spike IgG (G), anti-RBD IgG (H) and neutralizing Ig (I) before vaccination, before second dose administration and 1 and 6 months following the complete vaccination cycle in 86 naïve and 39 recovered COVID-19 subjects. Black asterisks represent unpaired statistic between naïve and recovered COVID-19 subjects at each time point calculated with Mann-Whitney test. * = *p*<0.05; ** = *p*<0.01; ***, ### = *p*<0.001.

Finally, recovered COVID-19 subjects before vaccination displayed significantly higher levels of anti-nucleoprotein (N) IgG than naïve individuals, confirming their previous exposure to SARS-CoV-2 (Figure 1F). These antibodies decreased over time reaching, at months 6 and 8, values below the cut-off limit. This finding confirms that also natural immunization to SARS-CoV-2 is subjected to decline over time. Naïve subjects instead exhibited anti-N IgG below cut-off value at all time points (Figure 1F).

Data obtained on an extended cohort of 86 naïve and 39 recovered COVID-19 subjects confirmed that anti-spike IgG, anti-RBD IgG and neutralizing Ig increase rapidly after the first vaccine injection in SARS-CoV-2 experienced individuals and then decline slowly up to month 6 post vaccination (Fig, 1G-I). On the contrary, in naïve subjects these antibodies peak after the second dose administration, at month 1 post vaccination, but then sharply decline at month 6 (Fig, 1G-I). Notably, antibody levels in naïve subjects did not reach the levels observed in recovered COVID-19 at all the analysed time points (Fig, 1G-I).

### Naïve and COVID-19 recovered subjects display comparable frequencies of spike-specific B cells at 8 months after vaccination

To further support the data obtained from the analysis of serum levels of SARS-CoV-2 spike-specific antibodies, we evaluated by flow cytometry the frequency of B cells expressing a B cell receptor (BCR) capable of recognizing SARS-CoV-2 spike protein (Fig. 2A). As shown in Fig.2B, in naïve subjects there was a gradual and significant increase in the frequency of B cells (CD19+) expressing a spike specific BCR (spike+) up to month 8 following the administration of the second dose of vaccine (left panel). On the contrary, in recovered-COVID-19 subjects, the significant increase in the frequency of spike-specific B cells observed following the first vaccine injection was followed by a progressive significant decline (middle panel). Of note, at month 8 both naïve and recovered COVID-19 subjects showed significantly higher levels of circulating spike-specific B cells than before vaccination. When comparing the frequency of these cells between the two study groups, we found that recovered COVID-19 subjects exhibited significantly higher frequencies of circulating spike-specific B cells from baseline to month 1, while at month 8 we observed comparable percentages (Fig.2B right panel). Circulating spike-specific B cells with a memory phenotype (CD27+) expressing on their surface a BCR of IgG, IgM or IgA isotype exhibited a similar kinetics to the one shown by total spike-specific B cells in both groups (Fig.2C-E left and middle panels). In addition, these cells were significantly more represented in recovered COVID-19 than in naïve subjects up to month 1 post vaccination, while exhibited similar frequencies at month 8 (Fig. 2C-E right panels).

**Figure 2.**
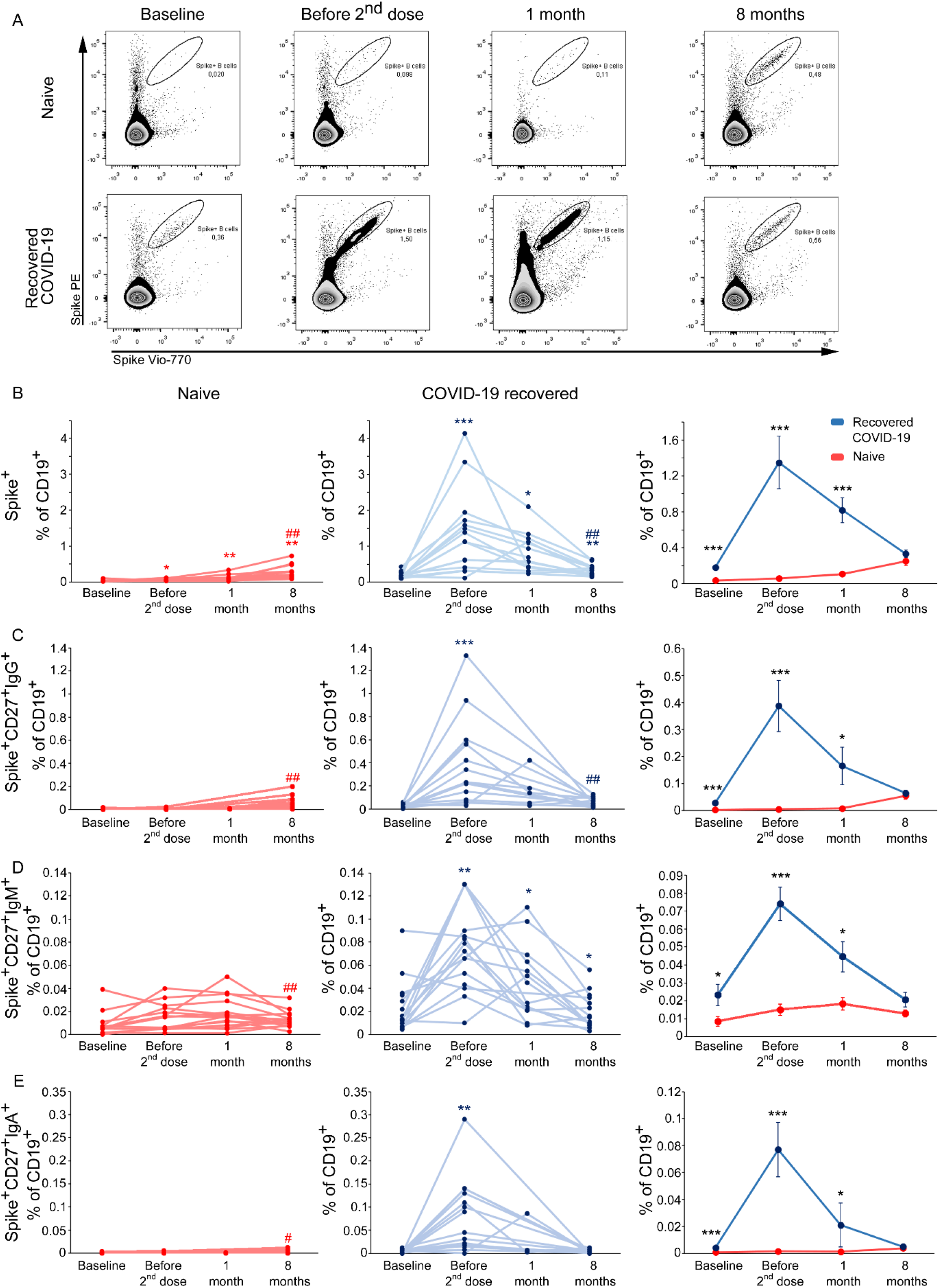
Longitudinal evaluation of spike-specific circulating B cells in naïve and COVID-19 recovered vaccinated subjects. (A) Representative flow cytometry plots of Spike-specific B cells in one naïve (upper row) and one COVID-19 recovered subject (lower row) before vaccination, before second injection and after 1 and 8 months following the complete vaccination cycle. Longitudinal evaluation of frequencies of total (B), CD27+ IgG+ (C), CD27+ IgM+ (D), CD27+ IgA+ (E) Spike-specific B cells in naïve (red lines, left column) and COVID-19 recovered (blue lines, middle column) subjects. Cumulative data are represented in right columns as mean ± SE from 15 naïve and 15 COVID-19 recovered subjects. Red and blue asterisks (left and middle columns) refer to paired statistics within each study group compared to the previous time point in the kinetic calculated with Wilcoxon-Signed Rank test. Red and blue hashtag in left and middle columns refer to paired statistics within each study group between baseline and 8 months calculated with Wilcoxon-Signed Rank test. Black asterisks in right column represent unpaired statistic between naïve and recovered COVID-19 subjects at each time point calculated with Mann-Whitney test. *, # = *p*<0.05; **, ## = *p*<0.01; *** = *p*<0.001.

### Frequency of spike-specific T cells is comparable in naïve and recovered COVID-19 subjects at month 1 post vaccination and it is stable up to month 8

To obtain a complete picture of the adaptive immune response to anti-COVID-19 vaccination, we evaluated the antigen-specific CD4+ T cell response to SARS-CoV-2 spike protein peptides, monitoring CD154 expression and the production of IL-2, IFN-γ and TNF-α upon in vitro stimulation (Fig. 3A). Total spike specific CD4+ T cells, defined by CD154 expression and the production of at least one among the three analysed cytokines, showed a gradual increase in naïve subjects until month 1 after the second vaccine dose, followed by a slight decrease at month 8 (Fig.3 B, left panel). On the contrary, in recovered-COVID-19 subjects, after an initial significant increase following the first vaccine dose, the frequency of these cells declined significantly at month 1 and then stabilized up to month 8 (middle panel). As observed for B cells, spike-reactive CD4+ T cells in both groups showed significantly higher frequencies at month 8 than before vaccination. In addition, total spike-specific CD4+ T cells were significantly higher in recovered COVID-19 than in naïve subjects until the administration of the second vaccine dose, while they became comparable at month 1 and remained similar also at month 8 (right panel). These observations were confirmed also when focusing on CD4+ T cells secreting specifically IFN-γ (Fig.3C), IL-2 (Fig.3D) or TNF-α (Fig.3E).

**Figure 3.**
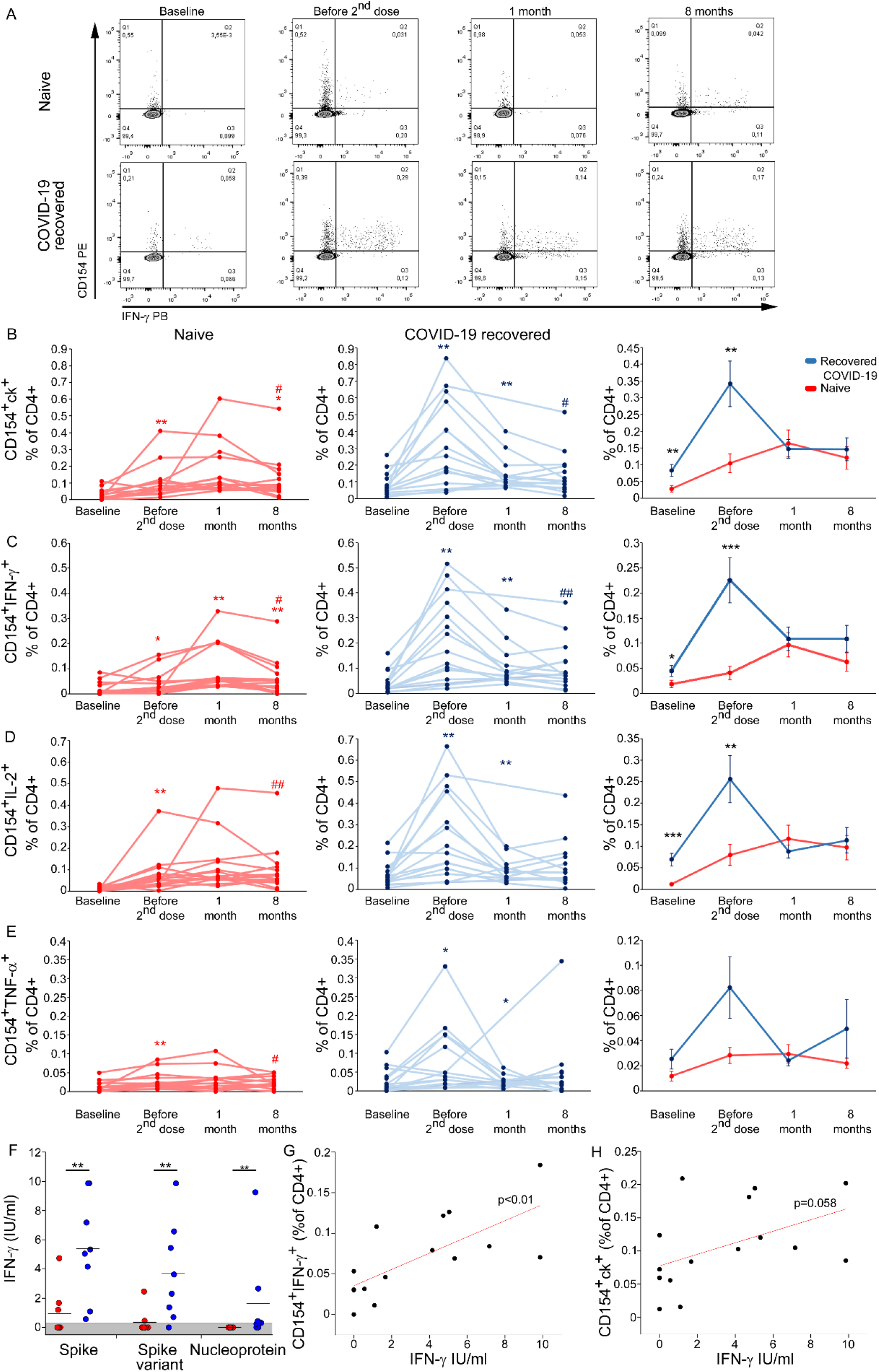
Longitudinal evaluation of spike-specific circulating T cells in naïve and COVID-19 recovered vaccinated subjects. A) Representative flow cytometric plots of Spike-specific CD4+ CD154+ IFN-γ+ T cells in one naïve (upper row) and one COVID-19 recovered (lower row) subject before vaccination, before second injection and after 1 and 8 months following the complete vaccination cycle. Longitudinal evaluation of frequencies of Spike-specific T cells expressing CD154 and producing at least one cytokine among IL-2, IFN-γ and TNF-α (B), CD154+ IFN-γ+ (C), CD154+ IL-2+ (D), CD154+ TNF-α+ (E) in naïve (red lines, left column) and COVID-19 recovered (blue lines, middle column) subjects. Cumulative data are represented in right columns as mean ±SE from 15 naïve and 15 COVID-19 recovered subjects. In panels B-E, represented data are subtracted of background, unstimulated condition. Red and blue asterisks (left and middle columns) refer to paired statistics within each study group compared to the previous time point in the kinetic calculated with Wilcoxon-Signed Rank test. Red and blue hashtag in left and middle columns refer to paired statistics within each study group between baseline and 8 months, calculated with Wilcoxon-Signed Rank test. Black asterisks in right column represent unpaired statistic between naïve and recovered COVID-19 subjects at each time point calculated with Mann-Whitney test. (F) Levels of IFN-γ in sera following whole blood stimulation with Spike, Spike variant (from Beta and Gamma variants) or Nucleoprotein performed on 8 naïve and 8 recovered COVID-19 subjects at month 8 post vaccination. Represented data are subtracted of background, unstimulated condition. (G) Correlation between the frequency of circulating CD4+CD154+IFN-γ+ cells and IFN-γ serum levels following whole stimulation with Spike. (H) Correlation between the frequency of circulating CD4+CD154+ cells producing at least one cytokine (among IFN-γ, IL-2 and TNF-α) and IFN-γ serum levels following whole blood stimulation with Spike. *, # = *p*<0.05; **, ## = *p*<0.01; *** = *p*<0.001.

To characterize more deeply the effector capacity of vaccine-induced CD4+ T cells, we also monitored the amount of IFN-γ secreted by cells collected at month 8 post vaccination and stimulated overnight with SARS-CoV-2 antigens. T cells from recovered COVID-19 subjects produced significantly higher levels of IFN-γ following stimulation with spike, if compared to naïve individuals (Figure 3F). Of note, the same difference was observed even when stimulating with spike variants (Beta and Gamma) (Figure 3F). As expected, IFN-γ was detected only in recovered COVID-19 individuals following stimulation with nucleoprotein, although in only 5 out of 8 tested subjects (Figure 3F). Interestingly, IFN-γ levels detected in the serum following spike peptide pool stimulation directly correlated with the frequency of CD154+IFN-γ+ spike-specific CD4+ T cells (Figure 3G). The strength of this correlation appeared partially reduced when correlating IFN-γ production with the frequency of total spike-specific CD4+ T cells, suggesting that IFN-γ secretion is not a peculiar feature of all spike-specific CD4+ T cells (Fig. 3H).

### Booster vaccination reactivates humoral and cellular immunity in naïve individuals more efficiently than in COVID-19 recovered subjects

The progressive decline in vaccine efficacy observed 6 months after the second dose administration led many countries to the decision to proceed with the administration of a third dose of vaccine (booster dose). In this regard, we had the opportunity to evaluate both humoral, B and CD4+ T cell-mediated spike-specific immune response in 14 (7 naïve and 7 ex-COVID-19) subjects before and one week after the administration of the booster dose. As shown in Fig.4A-B, serum titers of anti-S IgG and anti-RBD IgG significantly increased in naïve, but not in recovered COVID-19 subjects after the booster dose. Interestingly, while serum titers of spike neutralizing Ig were not affected by the booster dose in naïve, a significant reduction was induced in recovered COVID-19 subjects (Fig.4C). Despite a significantly different amount of anti-S IgG, anti-RBD IgG and neutralizing Ig before the booster, the two study groups showed comparable amount of these antibodies one week after the injection. In agreement with antibody data, the frequencies of spike-specific B cells and CD4+ T cells significantly increased in naïve, but not in recovered-COVID-19 subjects after the booster administration (Fig. 4D and E). In this case as well, spike-specific B and CD4+ T cells exhibited comparable frequencies between the two groups after booster.

**Figure 4.**
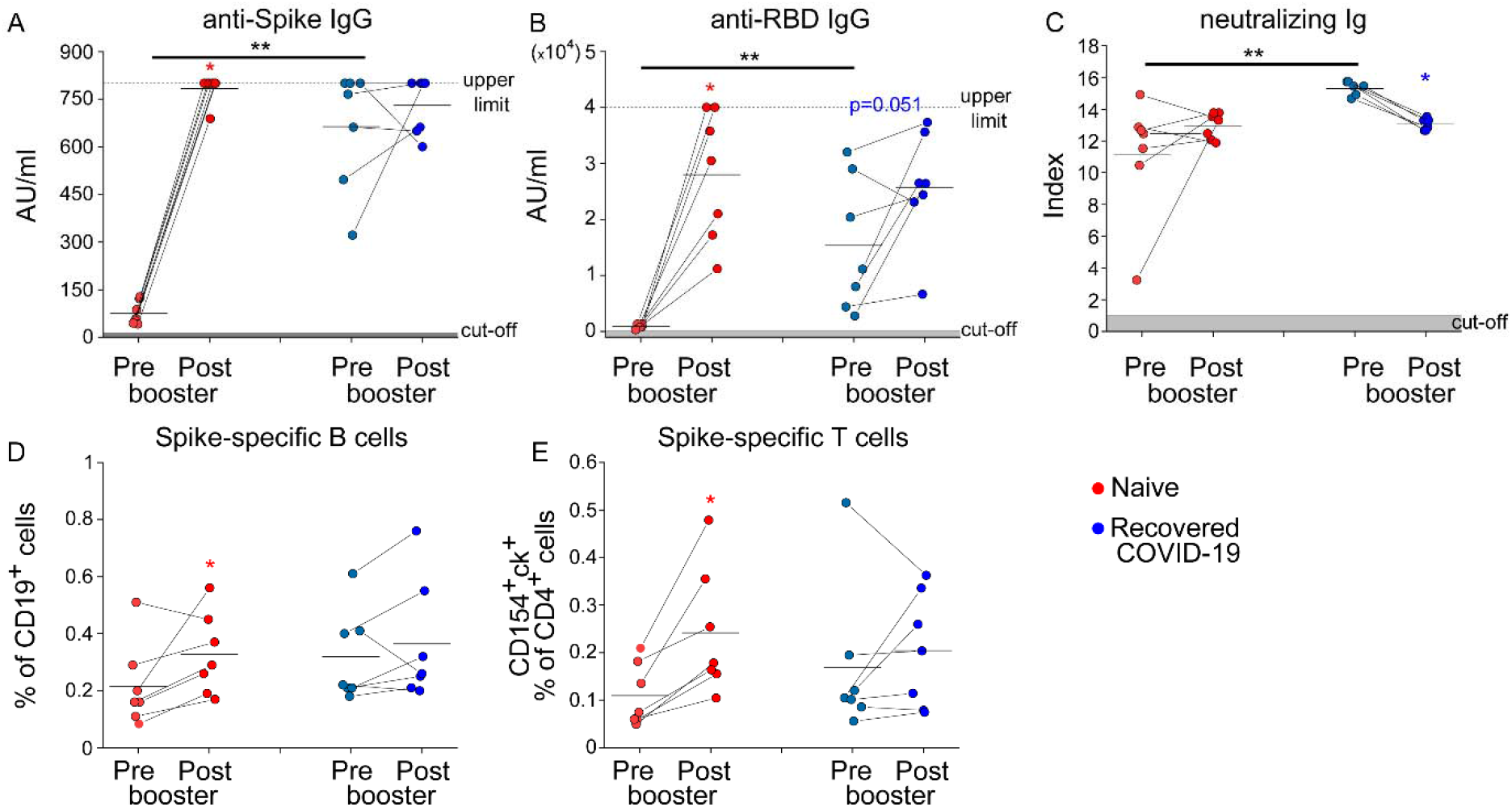
Reactivation of humoral and cellular immunity following vaccine booster administration in naïve and COVID-19 recovered subjects. Levels of anti-spike IgG (A), anti-RBD IgG (B), neutralizing Ig (C) in 7 naïve (red) and 7 recovered COVID-19 (blue) subjects before (pre) and 1 week after (post) vaccine booster injection. Levels of spike-specific B (D) and T (defined as CD154+ and producing at least one cytokine among IFN-γ+, IL-2 and TNF-α cells) in 7 naïve (red) and 7 recovered COVID-19 (blue) subjects before (pre) and 1 week after (post) vaccine booster injection. Red and blue asterisks refer to paired statistics within each study calculated with Wilcoxon-Signed Rank test. Black asterisks in right column represent unpaired statistic between naïve and recovered COVID-19 subjects calculated with Mann-Whitney test. * = *p*<0.05; ** = *p*<0.01

### Humoral, but not cellular immunity, declines up to one year following natural infection

In order to compare the longevity of immunological memory induced by vaccination or SARS-CoV-2 natural infection, we enrolled 14 recovered-COVID-19 subjects who were longitudinally sampled at 1, 6 and 12 months following hospital discharge. Since vaccine administration is recommended also in recovered COVID-19 individuals, this is a rather unique cohort as they were sampled before vaccine administration. We evaluated serum levels of anti-nucleoprotein IgG, anti-S IgM and IgG, anti-RBD IgG, spike neutralizing Ig, as well as the frequencies of spike-specific B and CD4+ T cells. As shown in Fig. 5, anti-nucleoprotein IgG, anti-S IgM and IgG, and anti-RBD IgG significantly declined at month 6 and even more at month 12 after hospital discharge (Fig.5A-D), whereas spike neutralizing Ig levels remained stable over time (Fig. 5 E).

**Figure 5.**
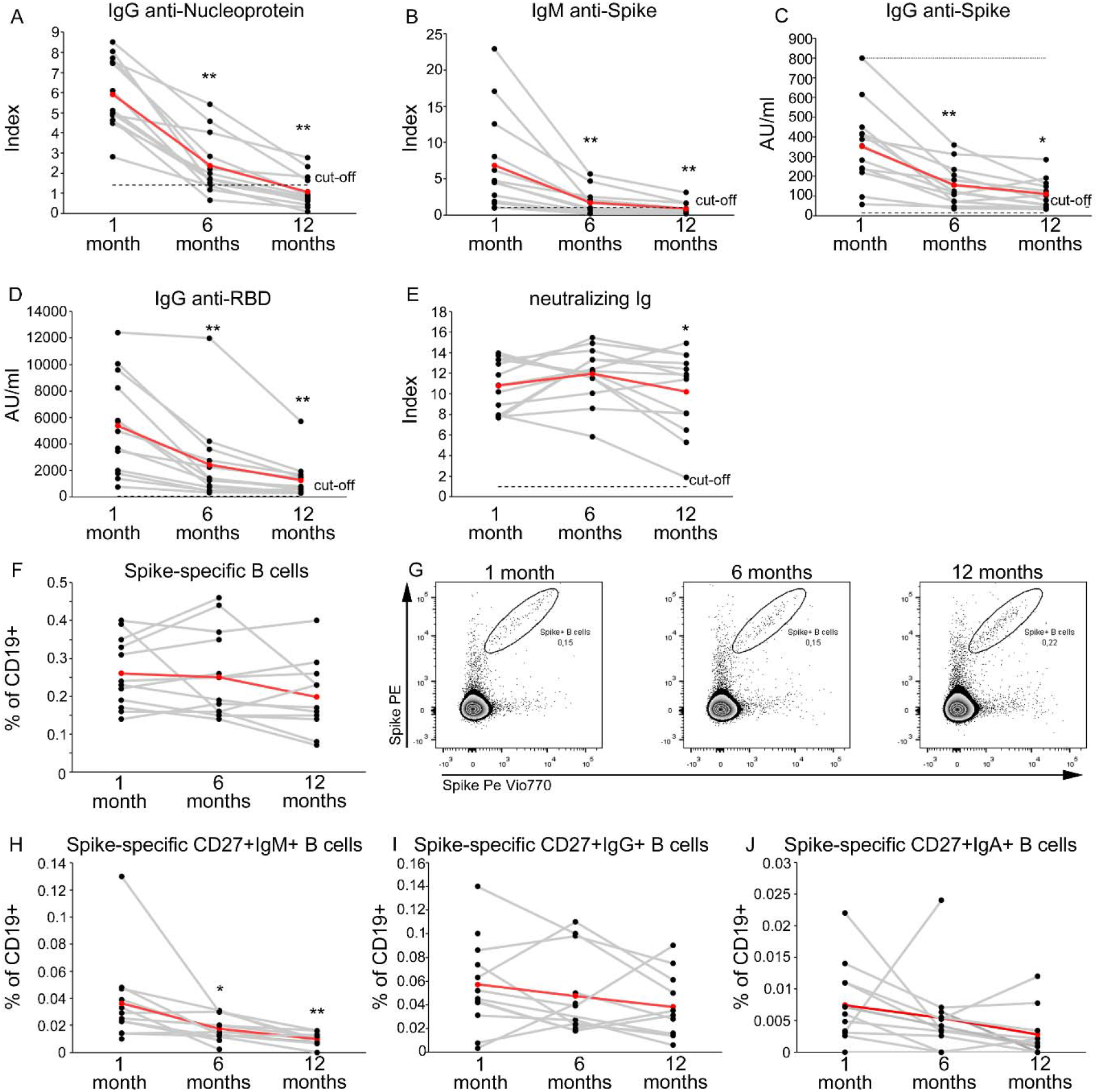
Longitudinal evaluation of SARS-CoV-2 specific antibody levels up to one year following infection. Levels of anti-N IgG (A), anti-S IgM (B), anti-S IgG (C), anti-RBD IgG (D), neutralizing Ig (E) in 13 recovered COVID-19 subjects evaluated 1, 6 and 12 months following hospital discharge. Frequencies of total (F), CD27+IgM+ (H), CD27+IgG+ (I), CD27+IgA+ (J) spike-specific B cells evaluated at 1, 6 and 12 months following hospital discharge in 14 recovered COVID-19 subjects. Representative FACS plots of total spike-specific B cells at each time point are shown in (G). Red lines represent mean values. Asterisks refer to paired statistics at each time point compared to the previous, calculated with Wilcoxon-Signed Rank test. * = *p*<0.05; ** = *p*<0.01

Prominently, the frequency of circulating B cells expressing a BCR capable of recognizing SARS-CoV-2 spike protein was substantially comparable over time (Fig.5F-G). Moreover, with the exception of spike-specific CD27+ IgM B cells, also the frequencies of CD27+ IgG+ and CD27+ IgA+ spike-specific B cells did not decline (Fig.5H-J). Similarly, the frequency of SARS-CoV-2-specific CD4+ T cells, as assessed by the expression of CD154 and the ability to produce at least one among IL-2, IFN-γ or TNF-α upon in vitro stimulation with spike or combination of spike, nucleoprotein and membrane peptides pools, was characterized by stability over time (Fig.6A-B and Fig.6F-G). The same stability was observed even when evaluating the production of specific cytokines (IFN-γ, IL-2 or TNF-α), in response to spike or to the combination of spike, membrane and nucleoprotein (Fig.6C-E and Fig.6H-J).

**Figure 6.**
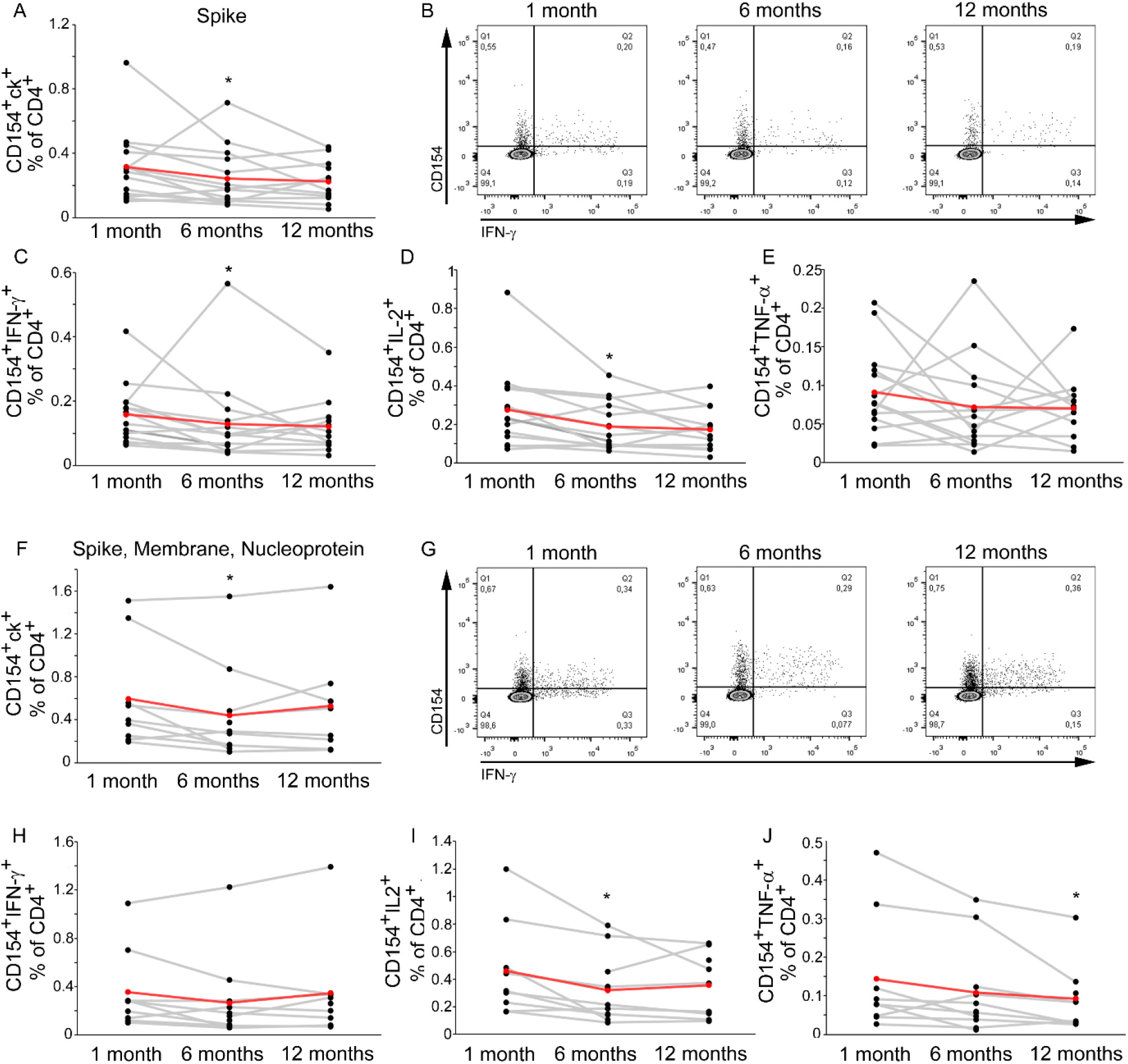
Longitudinal evaluation of SARS-CoV-2 specific B and T cells up to one year following infection. Frequencies of spike-specific CD4+ T cells expressing CD154 and producing at least one cytokine among IL-2, IFN-γ and TNF-α (A), CD154+ IFN-γ+ (C), CD154+ IL-2+ (D), CD154+ TNF-α+ (E) evaluated at 1, 6 or 12 months following hospital discharge in 14 recovered COVID-19 subjects. Representative FACS plots of CD154+IFN-γ+ spike-specific CD4+ T cells a at each time point are shown in (B). Frequencies of CD4+ T cells reactive to spike, membrane and nucleoprotein expressing CD154 and producing at least one cytokine among IL-2, IFN-γ and TNF-α (F), CD154+ IFN-γ+ (I), CD154+ IL-2+ (I), CD154+ TNF-α+ (J) evaluated at 1, 6 or 12 months following hospital discharge in 14 recovered COVID-19 subjects. Representative FACS plots of CD154+IFN-γ+ CD4+ T cells reactive to spike, membrane, nucleoprotein at each time point are shown in (G). Red lines represent mean values. Asterisks refer to paired statistics at each time point compared to the previous, calculated with Wilcoxon-Signed Rank test. * = *p*<0.05; ** = *p*<0.01

## Discussion

Anti-COVID-19 vaccines have shown remarkable efficacy both in phase 3 clinical trials as well as in real world studies, demonstrating protection against severe disease and infection [12-14]. Vaccination leads to the development of humoral and cellular immunity against spike [3,10]. However, it is currently unknown how long immunological memory induced by vaccination will last, with fundamental implications on vaccination strategies and the future course of the current pandemic. Epidemiological data have shown that 6 months following the complete two-doses mRNA vaccination cycle, vaccine effectiveness against infection drops down from 88% to 47% [7]. Nonetheless, protection against severe disease remains high when compared to unvaccinated people [8]. The reduced efficacy over time may be the result of the progressive waning of immunological memory, the occurrence of new SARS-CoV-2 variants with increased transmissibility and immune evasion potential, or a combination of both. In this study, we investigated the longevity of vaccine-induced humoral and cellular immunity to spike up to 8 months following vaccination in naïve and recovered COVID-19 individuals. We and others have previously shown that in recovered COVID-19 subjects one dose of mRNA vaccine is sufficient to achieve high levels of anti-spike immunity, higher than those observed in naïve subjects after two vaccine injections [3,8]. In line with this observation, we found here that anti-spike antibodies are maintained at higher levels in recovered COVID-19 individuals at months 6 and 8 post vaccination. Indeed, anti-spike IgG and anti-RBD IgG showed a significant drop from month 1 to month 6 in naïve subjects. Interestingly, neutralizing antibodies declined less rapidly in both groups. This peculiar kinetics could be explained by the fact that affinity maturation occurs over time. These findings are in agreement with a recent publication, showing that neutralizing antibodies decline slower than total anti-spike Ig in both naïve and recovered COVID-19 subjects [15].

Differently from antibody levels, we observed that anti-spike cellular immunity is comparable at month 8 between naïve and recovered COVID-19 individuals. Regarding spike-specific B cells, we found that the initial massive expansion observed early after the first vaccination in recovered COVID-19 subjects is followed by a significant contraction phase. On the contrary, in naïve individuals spike-specific B cells are very rare at initial time points, but then increase over time from month 1 to month 8. Spike-specific CD4+ T cells instead show comparable frequency in the two groups already at month 1 and then slowly decline in parallel up to month 8. Regarding the effector capacity, we observed that despite similar frequencies of vaccine-induced CD4+ T cells at month 8, recovered-COVID-19 subjects produced significantly more IFN-γ following stimulation with spike than naïve individuals. Of note, this observation was confirmed also when stimulating with SARS-CoV-2 spike from Beta and Gamma variants, confirming previous data showing that vaccine-induced T cell response is largely conserved against viral variants [16,17 and our observations under review]. It should be noted that the frequencies of spike-specific B and CD4+ T cells at month 8 are at least comparable to those observed in recovered COVID-19 subjects before vaccination, confirming that they are still detectable at this time point. These kinetics are similar to those observed by Goel et al., who monitored the persistence of spike-specific cellular immunity up to 6 months following vaccination [15]. The picture that comes out from these observations shows that antibody levels decline rapidly few months after vaccination, while memory B and CD4+ T cells display features of long-term persistence. This scenario is compatible with the loss of vaccine efficacy observed in real world studies 6 months after vaccination [7]. Indeed, the progressive waning of antibody levels coupled with the reduced neutralization efficacy of vaccinees’ sera against Delta and Omicron variant may expose to an increased risk of disease [18-21]. However, the presence of memory B and CD4+ T cells, which can rapidly reactivate following the encounter with SARS-CoV-2, can be a reason for the retained efficacy against severe disease. The administration of a third (booster) vaccine injection has shown to restore protection against symptomatic disease [22,23]. For this reason, given the increased numbers of breakthrough infections, several countries have initiated booster campaigns. We monitored the effect of a third dose injection in a cohort of naïve and SARS-CoV-2 experienced subjects. Our results show that the booster dose significantly increases the levels of spike-specific antibodies, B and CD4+ T cells in naïve individuals. This observation provides the immunological reason for the restored efficacy against symptomatic COVID-19. Longitudinal studies are needed to understand if a booster injection will lead to sustained maintenance of anti-spike humoral immunity, or whether a similar decline will occur in the coming months. In this case, it is plausible that regular boosters may be needed to keep vaccine efficacy high. The observation that SARS-CoV-2 infection followed by two vaccine injections results in longer maintenance of anti-spike antibodies, suggests that the third vaccine dose may result in longer antibody persistence in those unexposed to SARS-CoV-2. Differently from what observed in naïve individuals, the third vaccine dose has only minor effects on COVID-19 recovered subjects, in line with what observed for the second vaccine dose [3,10]. Given that at month 8 these individuals display high anti-spike antibody levels, as well as presence of memory B and CD4+ T cells, regulatory mechanisms may be involved in preventing an over-activation of anti-spike immunity. On the other hand, it may be possible that spike-specific B and CD4+ T cells display an exhausted phenotype, which prevents optimal reactivation upon antigen restimulation. Additional studies are needed to fully understand this phenomenon. However, this observation raises the question about the need for boosters in recovered COVID-19 subjects.

In this manuscript, we also monitored the long-term persistence of immunity to SARS-CoV-2 up to one year following infection in a group of moderate to severe COVID-19 subjects. This is a unique cohort, as in the majority of countries vaccination has been recommended also to individuals who have a history of SARS-CoV-2 infection. The data obtained are largely comparable to those observed following vaccination, with a progressive decline in antibody levels. Anti-nucleoprotein IgG display the shortest persistence, as their levels drop below the cut-off value at month 12. Notably, also in the case of natural infection, neutralizing antibodies decay slower than total anti-spike IgG and anti-RBD IgG. The progressive reduction of antibody levels may be the reason for the observed reinfections [24, 25]. On the contrary, the frequencies of spike-specific B and CD4+ T cells are largely stable up to one year from hospital discharge. The same stability was observed also when expanding our analysis to other SARS-CoV-2 antigens, as the frequency of CD4+ T cells reactive to spike, membrane and nucleoprotein was basically stable from month 1 to month 12. Collectively, these findings suggest that people recovering from moderate to severe COVID-19 may develop long-term persistent cellular immunity to SARS-CoV-2, in agreement with previous observations obtained on the original SARS-CoV showing that memory cells can be detected up to 17 years post infection [26]. This conclusion may not be extended to all SARS-CoV-2 infected subjects, as previous data have shown that asymptomatic infection leads to the development of reduced levels of immunological memory to the virus [5, 27].

In conclusion, our data show that vaccine-and infection-induced immunological memory display a similar kinetics of decay. Short-term persistence of humoral immunity may be responsible of reinfections, although long-lived memory B and CD4+ T cells may protect from severe disease development. A booster dose restores optimal anti-spike immunity in naïve subjects, while the need for vaccinated COVID-19 recovered subjects has yet to be defined, especially in those with previous moderate to severe disease. Epidemiological data may help to resolve this issue.

## Materials and Methods

### Subjects

30 healthcare workers who received BNT162b2 mRNA COVID-19 vaccine were recruited at the Careggi University Hospital, Florence, Tuscany, Italy, by the Infectious and Tropical Diseases Unit. Among them, 15 had a previous history of symptomatic SARS-CoV-2 infection (recovered COVID-19). Confirmation of SARS-CoV-2 infection was obtained by PCR analysis of nasopharyngeal swab. Negative history for infection in naïve individuals was based on absence of symptoms, absence of anti-N and anti-S IgG before receiving vaccination and routine monitoring by nasopharyngeal swab PCR testing. Among recovered COVID-19 individuals, disease severity was defined according to WHO guidelines [27]. Basic demographic and clinical characteristics are summarized in Supplementary Table S1. Following immunization schedule originally approved by the European Medicines Agency, each subject received two vaccine injections, 21 days apart. Blood samples were collected before the first dose (basal time, T0), before the second dose, and then 1 month, 6 months and 8 months later. 95 additional individuals (71 naïve and 24 COVID-19 recovered) were also recruited in addition to the 30 aforementioned subjects to expand serological data on a larger cohort. These subjects (125 in total, 86 naïve and 39 recovered COVID-19) were serologically tested before the first injection, before the second and then 1 month and 6 months later. Recruited subjects were not affected by chronic medical conditions that may affect vaccine response, with the exception of only one individual in the COVID-19 recovered group who was under immune suppressive treatment following solid organ transplantation. Basic demographic and clinical characteristics of the expanded cohort are summarized in Supplementary Table S2. 14 individuals, 7 of whom with previous SARS-CoV-2 infection, were sampled before booster BNT162b2 injection and one week after. All individuals received the booster dose 9 months after the second vaccine dose. Basic demographic and clinical characteristics are summarized in Supplementary Table S3.

Peripheral blood samples from 14 subjects hospitalized for COVID-19 between March and April 2020 were obtained 1 month, 6 months and 12 months following discharge. These individuals were not vaccinated at the last follow up date. Basic demographic and clinical characteristics are summarized in Supplementary Table S4.

PBMNCs were obtained following density gradient centrifugation of blood samples using Lymphoprep (Axis Shield Poc As(tm)) and were frozen in FCS plus 10% DMSO to be stored in liquid nitrogen. For each subject, longitudinal samples were defrosted and analyzed together for B cells and T cells evaluation. Serum was frozen and stored for Ig levels evaluation.

### Evaluation of SARS-CoV-2-Spike-reactive T cells

For T cells stimulation in vitro, 1.5 million PBMNCs were cultured in complete RPMI plus 5% human AB serum in 96 well flat bottom plates in presence of medium alone (background, negative control) or of a pool of Spike SARS-CoV-2 peptide pools (Prot_S1, Prot_S+ and Prot_S to achieve a complete sequence coverage of the Spike protein) at 0.6 µM/peptide, accordingly to manufacturer’s instructions (Miltenyi Biotech). Cells from recovered COVID-19 subjects longitudinally assessed up to 1 year following hospital discharge were additionally stimulated with peptide pools of Membrane protein and Nucleoprotein from SARS-CoV-2 at 0.6 µM/peptide, accordingly to manufacturer’s instructions (Miltenyi Biotech). After 2 hours of incubation at 37°C, 5% CO2, Brefeldin A (5 µg/mL) was added, followed by additional 4 hours incubation. Finally, cells were fixed and stained using fluorochrome-conjugated antibodies listed in Table S5. Samples were acquired on a BD LSR II flow cytometer (BD Biosciences) with FACSDiva Software and analysed by FlowJo Software. The gating strategy applied has already been described [3,4].

### Evaluation of SARS-CoV-2 spike-specific B cells

For B cells evaluation, 2 million PBMNCs were stained for 30 minutes at 4°C with fluorochrome-conjugated antibodies listed in Table S6, then washed with PBS/EDTA buffer (PEB), and incubated 5 minutes with 7-AAD for viability evaluation. Recombinant biotinylated SARS-Cov2 Spike protein (Miltenyi Biotech) was conjugated separately with streptavidin PE and PE-Cy7 for 15 minutes at room temperature and pooled in 1:2 ratio, before being added to final staining mix. Samples were acquired on a BD LSR II flow cytometer (BD Biosciences) with FACSDiva Software and analysed by FlowJo Software. The gating strategy applied has already been described [3].

### Evaluation of SARS-CoV-2-specific IgM and IgG

Evaluation of SARS-CoV-2 Spike protein antibodies, including anti-Spike (in trimeric form)-specific IgG (Diasorin), anti-Spike RBD-specific IgG (Abbott) and IgA (Euroimmun), anti-Spike-specific IgM (Abbot), anti-Nucleoprotein-specific IgG (Abbott), and neutralizing antibodies which block binding of Spike protein with the ACE2 receptor (Dia.Pro Diagnostic Bioprobes) was performed following manufacturers’ instructions. The antibody reactivity of each specimen was expressed in AU/ml, or by the ratio between optical density and cut-off value (index).

### Evaluation of IFN-γ in culture supernatants following blood stimulation with SARS-CoV-2 antigens

Evaluation of IFN—γ in culture supernatants following blood stimulation with SARS-CoV-2 antigens was performed with STANDARDTM F Covi-FERON FIA (IFN-gamma) SD Biosensor according to the manufacturer’s instructions. Viral antigens used for stimulation included SARS-CoV-2 Spike Protein (SP) antigens derived from original SARS-CoV-2, and the 20I/501Y.V1 (Alpha), 20H/501 (Beta) and 20J/501Y.V3 (Gamma) viral variants, and antigens derived from the Nucleocapsid Protein (NP). The results were expressed as IU/ml of IFN-γ, subtracted of unstimulated background conditions.

### Statistical analysis

Unpaired Mann-Whitney test was used to compare COVID-19 recovered subjects versus naïve subjects; Wilcoxon-Signed Rank test was used to compare different time points in each study group. In all cases, p values ≤0.05 were considered significant.

### Study approval

The procedures followed in the study were approved by the Careggi University Hospital Ethical Committee. Written informed consent was obtained from recruited patients.

## Data Availability

All data produced in the present work are contained in the manuscript

## Acknowledgments

We thank all the subjects who participated to the study. This study was supported by funds to the Department of Experimental and Clinical Medicine, University of Florence (Project Excellence Departments 2018-2022), by the University of Florence, project RICTD2122, by the Italian Ministry of Health (COVID-2020-12371849) and by Tuscany Region (TagSARS CoV 2).

## Author Contributions

G.M.R., A.B., F.A., designed the study; N.D.L., E.M., M.S., L.Z. F.L. collected peripheral blood samples and informed consent; F.L., L.C. provided advice; A.M., L.M., L.S., A.V., M.C., G.L., S.T.K., M.G.C., A.R., A.C. P.F., performed experiments; A.M., L.M., analyzed data; A.M., L.M., F.A. wrote the manuscript. All authors revised the manuscript and gave final approval.

## Notes

**Conflict of interest statement:** The authors have declared that no conflict of interest exists.

### Competing Interest Statement

The authors have declared no competing interest.

### Author Declarations

Ethics committee of Area Vasta Toscana Centro gave ethical approval for this work

