## Supplementary data for "Long-lasting cellular immunity to SARS-CoV-2 following infection or vaccination and implications for booster strategies"

**Supplementary Table S1.** Basic demographic and clinical data of 30 healthcare workers who received BNT162b2 mRNA COVID-19 vaccine and were followed up to 8 months.

| | Subject ID | Gender | COVID-19 severity |
|---|---|---|---|
| **COVID-19 recovered individuals** | R1 | male | mild |
| | R2 | female | mild |
| | R3 | female | severe |
| | R4 | female | mild |
| | R5 | male | moderate |
| | R6 | male | mild |
| | R7 | female | mild |
| | R8 | male | moderate |
| | R9 | female | mild |
| | R10 | female | mild |
| | R11 | male | mild |
| | R12 | male | critical |
| | R13 | male | critical |
| | R14 | female | mild |
| | R15 | female | mild |
| Mean±SD | - | - | - |
| **Naïve individuals** | N1 | male | na |
| | N2 | female | na |
| | N3 | male | na |
| | N4 | female | na |
| | N5 | female | na |
| | N6 | female | na |
| | N7 | male | na |
| | N8 | female | na |
| | N9 | male | na |
| | N10 | male | na |
| | N11 | male | na |
| | N12 | female | na |
| | N13 | female | na |
| | N14 | female | na |
| | N15 | male | na |
| Mean±SD | - | - | - |

*na* denotes *not applicable*

**Supplementary Table S2.** Basic demographic and clinical data of 125 healthcare workers who received BNT162b2 mRNA COVID-19 vaccine and were followed up to 6 months.

| | **COVID-19 recovered individuals** | **Naïve individuals** |
|---|---|---|
| **No. subjects** | 39 | 86 |
| **Age** (years)<br>Mean±SD | 50.4±11.3 | 45.5±14.3 |
| **Gender**<br>- Male, n (%)<br>- Female, n (%) | <br>12 (31.6)<br>26 (68.4) | <br>39 (45.3)<br>47 (54.6) |
| **COVID-19 severity**<br>- Asymptomatic/mild, n (%)<br>- Moderate, n (%)<br>- Severe, n (%)<br>- Critical, n (%) | <br>31 (79.5)<br>2 (5.1)<br>2 (5.1)<br>4 (10.3) | Na |
| **Time from COVID-19 diagnosis° to first vaccine dose (days)**<br>- Mean±SD | 230.4±93.6 | na |

*na* denotes *not applicable*

**Supplementary Table S3.** Basic demographic and clinical data of 14 healthcare workers who received booster BNT162b2 mRNA COVID-19 vaccine injection and were followed up to one week.

| | **Subject ID** | **Gender** | **COVID-19 severity** |
|---|---|---|---|
| **COVID-19 recovered individuals receiving booster dose** | RB1 | female | severe |
| | RB2 | female | mild |
| | RB3 | male | critical |
| | RB4 | male | moderate |
| | RB5 | female | mild |
| | RB6 | male | critical |
| | RB7 | female | mild |
| Mean±SD | - | - | - |
| **Naïve individuals receiving booster dose** | NB1 | female | na |
| | NB2 | female | na |
| | NB3 | male | na |
| | NB4 | female | na |
| | NB5 | male | na |
| | NB6 | male | na |
| | NB7 | male | na |
| Mean±SD | - | - | - |

*na* denotes *not applicable*

**Supplementary Table S4.** Basic demographic and clinical data of 14 individuals who were hospitalized for COVID-19 between March and April 2020 and followed up to 12 months from discharge.

| | **Subject ID** | **Gender** | **COVID-19 severity** |
|---|---|---|---|
| **Unvaccinated COVID-19 recovered individuals** | U1 | male | moderate |
| | U2 | male | moderate |
| | U3 | male | severe |
| | U4 | female | severe |
| | U5 | male | moderate |
| | U6 | male | severe |
| | U7 | male | critical |
| | U8 | male | critical |
| | U9 | female | severe |
| | U10 | female | critical |
| | U11 | female | moderate |
| | U12 | female | moderate |
| | U13 | female | moderate |
| | U14 | male | critical |
| Mean±SD | - | - | - |

**Supplementary Table S5. List of all fluorochrome mAbs used for flow cytometric analysis of antigen specific T cells.**

| Antigen | Flurochrome | Clone | Company |
|---|---|---|---|
| TNF-α | FITC | 6401.1111 | BDBioscience |
| CD154 | PE | TRAP1 | BDBioscience |
| CD3 | PerCP | SK7 | BDBioscience |
| CD4 | PE-Cy7 | SK3 | Invitrogen |
| CD8 | Super Bright 600 | SK1 | eBioscience™ |
| IL-2 | APC | MQ1-17H12 | BDBioscience |
| IFN-γ | Pacific Blue | B27 | BioLegend |
| L/D | Fixable Viability Stain 780 | | BDBioscience |

**Supplementary Table S6. List of all fluorochrome mAbs used for flow cytometric analysis of antigen specific B cells.**

| **Antigen** | **Flurochrome** | **Clone** | **Company** |
|---|---|---|---|
| CD19 | APC-Vio770 | LT19 | Miltenyi |
| CD27 | VioBright FITC | M-T271 | Miltenyi |
| IgA | VioGreen | IS11-8E10 | Miltenyi |
| IgG | VioBlue | IS11-3B2.2.3 | Miltenyi |
| CD14 | PerCP | TÜK4 | Miltenyi |
| IgM | APC | PJ2-22H3 | Miltenyi |
| CD3 | PerCP | BW264/56 | Miltenyi |
| 7AAD | | | Miltenyi |
| Spike | PE | | Miltenyi |
| Spike | PE-Vio770 | | Miltenyi |
